# Seasonal Dynamics of Influenza and RSV in the Caribbean: A Call for Regionally Tailored Preventive Measures

**DOI:** 10.1101/2025.09.09.25335442

**Authors:** Charlene Maria, Jayant Kalpoe, Angelino Tromp, Fleur Koene, Dennis Souverein, Sonja van Roeden, Martijn Tilanus, Juldany Juliet, Marquita Euson, Sherryl Carty, Shanna Holaman, Sharda Baboe-Kalpoe, Fazal Baboe, Rianne Plaisier, Anneke Visser, Winny van Luling, Felix Holiday, Chérina Fleming, Josephine van de Maat, Peter Klein Klouwenberg, Radjinkoemar Steingrover, Lilly M. Verhagen

**Author notes:** Shared last authorship (Radjinkoemar Steingrover, Lilly M. Verhagen).

## Abstract

**Introduction:** Respiratory tract infections (RTIs) remain a leading global cause of morbidity and mortality, with the Caribbean reporting some of the highest incidence rates. The World Health Organization recommends tailoring prevention strategies to local viral epidemiology. We aim to characterize the seasonal trends and disease burden of major respiratory viruses in the Caribbean region of the Kingdom of the Netherlands.

**Methods:** We conducted a retrospective observational study using virological surveillance data routinely collected between 2018 and 2024 from Aruba, Bonaire, Curaçao, Sint Maarten, Saba, and Sint Eustatius. Seasonal patterns of rhinovirus, influenza virus and respiratory syncytial viruses (RSV) were modelled using generalised additive models. Associations with climate, tourism, age, and disease severity were assessed with generalised linear models.

**Results:** Rhinovirus was the most frequently detected virus across all islands. Influenza virus peaked between November and March (p < 0.001), aligning with seasonal trends in the Northern Hemisphere and coinciding with the high tourism season in Aruba (OR = 8.72; 95% CI: 6.37–12.10), Curaçao (OR = 3.05; 95% CI: 1.59–6.16), and Sint Maarten (OR = 10.83; 95% CI: 2.13–198.83). In contrast, RSV activity peaked from June to December (p < 0.001), corresponding with the rainy season in Aruba (OR = 6.42; 95% CI: 4.26–9.75), and Sint Maarten (OR = 7.27; 95% CI: 2.31–28.22). Rhinovirus detection was significantly associated with increased disease severity, including the need for oxygen therapy (OR = 2.78; 95% CI: 1.72-4.50) and presentation with dyspnoea or tachypnoea (OR = 2.26; 95% CI: 1.42-3.67).

**Conclusions:** RSV seasonality in the Caribbean aligns with the rainy season and diverged from patterns in the Netherlands, indicating that current European-based intervention schedules may not be optimally timed. By integrating virological surveillance data from all six islands, this study offers a unique regional perspective to inform public health policy.

## Introduction

Respiratory tract infections (RTIs), especially lower respiratory tract infections (LRTIs), remain a leading cause of death among communicable diseases worldwide. Children under the age of five, the elderly and immunocompromised individuals are particularly vulnerable.^1–6^ In 2021, LRTIs (excluding coronavirus disease 2019 (COVID-19)) were the fifth leading cause of death globally, claiming approximately 2.5 million lives.^1, 7^ Although age-standardised mortality rates for LRTIs declined by 48.5% between 1990 and 2019, this progress was reversed by COVID-19.^8^ According to the Global Burden of Disease Study 2021, global life expectancy declined by 1.6 years from 2019 to 2021, largely due to increased pandemic-related mortality.^7^ In the same year, Latin America and the Caribbean were disproportionately affected, recording the world’s highest age-standardised COVID-19 death rates and a life expectancy drop of 3.6 years.^7^ In 2019, the LRTI mortality rate in the Caribbean was 39.3 per 100,000, nearly three times higher than in Western Europe (13.7) and North America (13.2), highlighting persistent regional health inequities.^8^

RTIs are caused by a wide range of viral and bacterial pathogens.^2,5,9,10^ *Streptococcus pneumoniae* remains the leading bacterial cause in both children and adults. Among viruses, rhinovirus predominates in upper respiratory infections, while influenza virus is a major contributor to severe disease in older adults. Respiratory syncytial virus (RSV) is a leading cause of paediatric hospitalization, especially in children under the age of five. From 2019 to 2021, severe acute respiratory syndrome coronavirus 2 (SARS-CoV-2) emerged as the dominant RTI pathogen, surpassing all others.^5^ Clinical presentations vary widely, from mild upper respiratory symptoms to severe pneumonia and respiratory failure requiring hospitalization.

The World Health Organization (WHO) emphasises the importance to monitor local viral epidemiology to guide preventive measures such as vaccination and prophylaxis.^11,12^ However, for the six Caribbean islands of Aruba, Bonaire, Curaçao, Sint Maarten, Saba and Sint Eustatius, which are part of the Kingdom of the Netherlands, data on the seasonal circulation of key respiratory viruses, including influenza and RSV remain limited. Currently, these islands follow the temperate Dutch mainland schedule for influenza vaccination and RSV prophylaxis. Yet, their distinct geographic and climatic conditions suggest that transmission patterns may differ substantially.^13–16^ Improving our understanding of local viral seasonality is crucial for optimizing the timing of prevention strategies.^11^

Environmental and social factors, such as temperature, humidity, latitude, air quality and human behaviour, shape the timing and intensity of respiratory virus circulation.^13–16^ In temperate regions, influenza and RSV typically peak in winter, while rhinovirus circulates year-round with peaks in autumn.^13–16^ In contrast, tropical or subtropical regions often experience year-round influenza activity and RSV surges during the rainy season.^13–15^ Human mobility, particularly tourism, plays a key role in viral transmission.^17–19^ The six Caribbean islands attract tourists year-round from regions such as the Netherlands, the United States, Canada, Colombia and Brazil. Frequent inter-island travel further increases the potential for virus spread across the region. While island-specific data are limited, global travel is a well-established driver of respiratory virus dissemination.^17,20^ The COVID-19 pandemic underscored this link: international travel accelerated the spread of SARS-CoV-2 viral, while mobility restrictions and vaccination proved effective in curbing transmission.¹⁸⁻²¹

Despite the growing recognition of the role of climate and travel in respiratory virus transmission, their impact and the seasonal patterns of respiratory infections remain understudied in the Caribbean. Therefore, this study aims to (1) characterise the seasonal trends of viral RTIs in the Caribbean region of the Kingdom of the Netherlands; (2) assess the influence of climate and tourism on RTI prevalence, while taking into account the COVID-19 pandemic; (3) evaluate the regional disease burden associated with these infections. By addressing these objectives, this study aims to provide evidence-based insights to guide locally tailored public health interventions and optimise resource allocation for RTI prevention and control across the six islands.

## Methods

### Study Design

This retrospective observational study analyses the epidemiology of RTIs on the Caribbean islands of Aruba, Bonaire, Curaçao, Sint Maarten, Saba, and Sint Eustatius. Anonymised electronically available healthcare data were collected from diagnostic laboratories and hospitals from all six islands from patients tested for RTIs between January 2018 and September 2024. Cases were excluded if demographic data were incomplete or if testing was performed exclusively for travel-related purposes during the COVID-19 pandemic. The data from Sint Maarten, Saba, and Sint Eustatius included clinical information about the severity of disease that were used to assess the regional clinical burden of disease caused by viral RTIs. Exclusion criteria for this analysis included incomplete demographic data and patients who were tested exclusively for COVID-19 while under quarantine in the hospital.

### Data Collection

An anonymised dataset was compiled from all six islands, including diagnostic results for 8 respiratory viruses and their subtypes: adenovirus; endemic coronaviruses (229E, HKU1, NL63, OC43); SARS-CoV-2; human metapneumovirus (hMPV); rhinovirus/enterovirus; influenza A (subtypes A/H1, A/H3, A/H1-2009); influenza B; parainfluenza viruses types 1– 4; and respiratory syncytial virus types A and B (RSV A/B). For analyses, influenza A and B were combined and reported as “influenza”, and rhinovirus and enterovirus were combined and reported as “rhinovirus/enterovirus”.

Testing strategies varied both across and within islands over time, shaped by local clinical policies and resource availability. Diagnostics were performed using BIOFIRE® Respiratory and Pneumonia Panels, Seegene Allplex™ Respiratory Panels 1–4, and the Cepheid Xpert® Xpress CoV-2/Flu/RSV Plus assay. While SARS-CoV-2 antigen tests were widely used outside of clinical laboratories, particularly in public health and travel-related settings, most of these results were not captured in healthcare information systems and were therefore excluded. All diagnostic Polymerase Chain Reaction (PCR) results were included in the final dataset and aggregated for analysis.

Clinical data was available for patients hospitalised at the Sint Maarten Medical Center (SMMC), the general hospital for patients from Sint Maarten, Saba, and Sint Eustatius. Clinical variables included presenting symptoms, underlying comorbidities, and indicators of disease severity. Severity indicators comprised oxygen therapy requirements, presence of dyspnoea or tachypnoea, intensive care unit (ICU) admission, and length of hospital stay. Common symptoms such as coughing, sneezing, and fever were documented. Comorbidity profiles were also assessed, with the most frequently observed conditions identified.

### Analysis of Seasonal Trends

Monthly viral detection rates were analysed to identify peak infection periods. To better capture typical seasonal dynamics under stable conditions, data from the peak of the COVID-19 pandemic (2020-2021) were excluded from the primary analysis. Sensitivity analyses including this period were performed separately.

A complimentary analysis assessed the influence of climate variation, tourism influx and demographic variables (age and sex) on the seasonality of respiratory viruses. To account for shifts in viral circulation related to COVID-19, data were stratified into two periods: pre-/during-pandemic, and post-pandemic, using June 1, 2021, as the cut-off.

The Caribbean islands exhibit distinct climate patterns. Aruba, Bonaire and Curaçao have a semi-arid climate with a short rainy season from September to December, peaking in October.^21,22^ In contrast, Sint Maarten, Saba, and Sint Eustatius experience a tropical monsoon climate with a longer rainy season from June to December, also peaking in October.^23^ For analytical consistency across the entire region, and given the overlapping peak rainfall periods, the rainy season across the six islands was defined as June to December for the purpose of this study. This harmonised definition allows for standardised seasonal comparisons across islands with differing climate zones. High tourism season was defined as the period with peak tourist activity from December to April, and in July.^24–27^

### Statistical analysis

Absolute and relative numbers of tests and positive detections were calculated for each island. Descriptive statistics, including chi-square tests for categorical variables, were used to assess the differences in demographics, virus distribution, and seasonal patterns. To examine temporal trends in virus circulation, a time series analysis was conducted using generalised additive models (GAMs), with viral detection counts as the outcome variable. GAMs extend generalised linear models (GLMs) by incorporating nonlinear relationships through smoothing functions. Given the count-based nature of the outcome, a Poisson distribution was specified. GLMs were additionally used to evaluate the association between viral detections and climate, tourism, age group, and sex.

For the clinical dataset, descriptive statistics were used to summarise demographic characteristics, respiratory virus distribution, clinical symptoms, comorbidities, and indicators of disease severity. Group differences for these characteristics were assessed using chi-square tests for categorical variables and one-way analysis of variance (ANOVA) for continuous variables where appropriate. GLMs were applied to evaluate the association between respiratory viral detections and key indicators of disease severity.

All statistical analyses were performed in R (version 4.3.3), using the *tidyverse*, *mgcv*, *stats*, and *ggplot2* packages.

## Results

### Cohort description

A total of 35,647 tests for respiratory viruses were analysed across the six Caribbean islands, with 9,431 tests (26.5%) yielding positive results for at least one of the 8 target viruses. The cohort consisted predominantly of adults (n=26,656 tests, 74.8%; mean age = 38, IQR: 17– 59) with slightly more females (19,445 tests, 54.5%). However, on St Maarten and St Eustatius, the majority of patients were children (n=624, 58.5% and n=155, 50.8%, respectively; Table 1).

**Table 1.**
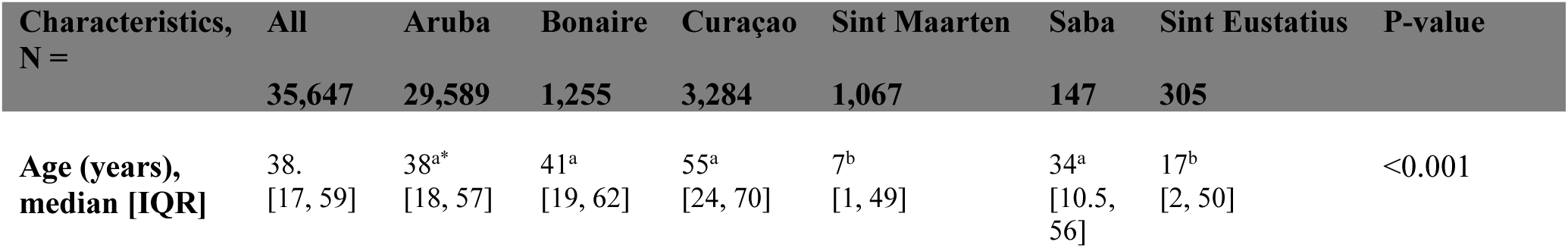
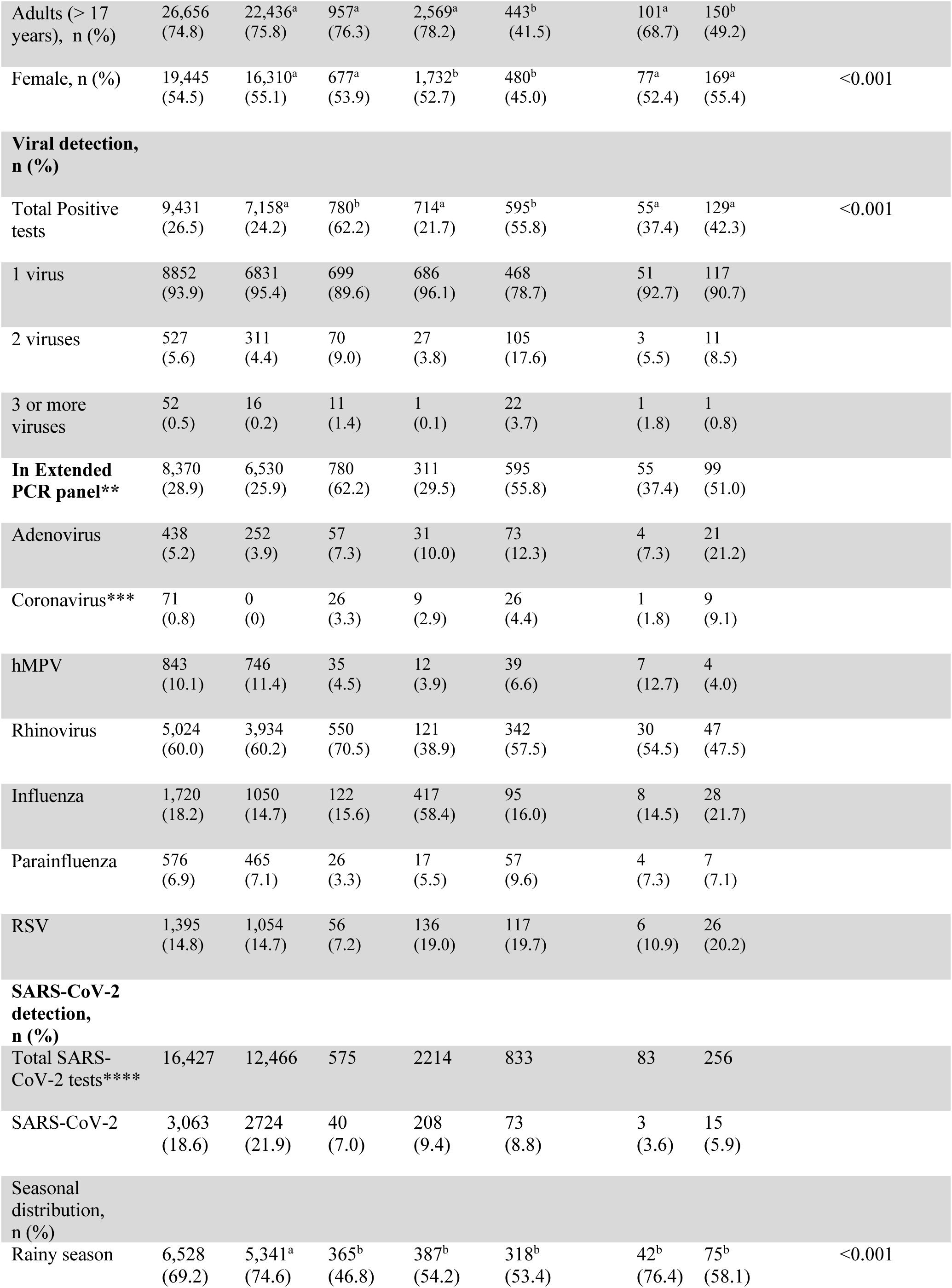
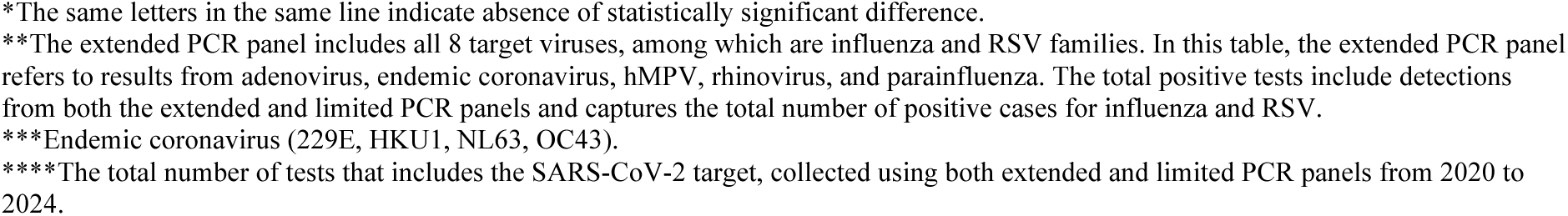
Demographics and Viral Detection Distribution.

| Characteristics,<br>N = | All<br>35,647 | Aruba<br>29,589 | Bonaire<br>1,255 | Curaçao<br>3,284 | Sint Maarten<br>1,067 | Saba<br>147 | Sint Eustatius<br>305 | P-value |
| --- | --- | --- | --- | --- | --- | --- | --- | --- |
| Age (years),<br>median [IQR] | 38.<br>[17, 59] | 38<sup>a*</sup><br>[18, 57] | 41<sup>a</sup><br>[19, 62] | 55<sup>a</sup><br>[24, 70] | 7<sup>b</sup><br>[1, 49] | 34<sup>a</sup><br>[10.5,<br>56] | 17<sup>b</sup><br>[2, 50] | <0.001 |
| Adults (> 17 years), n (%) | 26,656<br>(74.8) | 22,436<sup>a</sup><br>(75.8) | 957<sup>a</sup><br>(76.3) | 2,569<sup>a</sup><br>(78.2) | 443<sup>b</sup><br>(41.5) | 101<sup>a</sup><br>(68.7) | 150<sup>b</sup><br>(49.2) |  |
| Female, n (%) | 19,445<br>(54.5) | 16,310<sup>a</sup><br>(55.1) | 677<sup>a</sup><br>(53.9) | 1,732<sup>b</sup><br>(52.7) | 480<sup>b</sup><br>(45.0) | 77<sup>a</sup><br>(52.4) | 169<sup>a</sup><br>(55.4) | <0.001 |
| **Viral detection,**<br>**n (%)** |  |  |  |  |  |  |  |  |
| Total Positive tests | 9,431<br>(26.5) | 7,158<sup>a</sup><br>(24.2) | 780<sup>b</sup><br>(62.2) | 714<sup>a</sup><br>(21.7) | 595<sup>b</sup><br>(55.8) | 55<sup>a</sup><br>(37.4) | 129<sup>a</sup><br>(42.3) | <0.001 |
| 1 virus | 8852<br>(93.9) | 6831<br>(95.4) | 699<br>(89.6) | 686<br>(96.1) | 468<br>(78.7) | 51<br>(92.7) | 117<br>(90.7) |  |
| 2 viruses | 527<br>(5.6) | 311<br>(4.4) | 70<br>(9.0) | 27<br>(3.8) | 105<br>(17.6) | 3<br>(5.5) | 11<br>(8.5) |  |
| 3 or more viruses | 52<br>(0.5) | 16<br>(0.2) | 11<br>(1.4) | 1<br>(0.1) | 22<br>(3.7) | 1<br>(1.8) | 1<br>(0.8) |  |
| **In Extended PCR panel\*\*** | 8,370<br>(28.9) | 6,530<br>(25.9) | 780<br>(62.2) | 311<br>(29.5) | 595<br>(55.8) | 55<br>(37.4) | 99<br>(51.0) |  |
| Adenovirus | 438<br>(5.2) | 252<br>(3.9) | 57<br>(7.3) | 31<br>(10.0) | 73<br>(12.3) | 4<br>(7.3) | 21<br>(21.2) |  |
| Coronavirus*** | 71<br>(0.8) | 0<br>(0) | 26<br>(3.3) | 9<br>(2.9) | 26<br>(4.4) | 1<br>(1.8) | 9<br>(9.1) |  |
| hMPV | 843<br>(10.1) | 746<br>(11.4) | 35<br>(4.5) | 12<br>(3.9) | 39<br>(6.6) | 7<br>(12.7) | 4<br>(4.0) |  |
| Rhinovirus | 5,024<br>(60.0) | 3,934<br>(60.2) | 550<br>(70.5) | 121<br>(38.9) | 342<br>(57.5) | 30<br>(54.5) | 47<br>(47.5) |  |
| Influenza | 1,720<br>(18.2) | 1050<br>(14.7) | 122<br>(15.6) | 417<br>(58.4) | 95<br>(16.0) | 8<br>(14.5) | 28<br>(21.7) |  |
| Parainfluenza | 576<br>(6.9) | 465<br>(7.1) | 26<br>(3.3) | 17<br>(5.5) | 57<br>(9.6) | 4<br>(7.3) | 7<br>(7.1) |  |
| RSV | 1,395<br>(14.8) | 1,054<br>(14.7) | 56<br>(7.2) | 136<br>(19.0) | 117<br>(19.7) | 6<br>(10.9) | 26<br>(20.2) |  |
| **SARS-CoV-2**<br>**detection,**<br>**n (%)** |  |  |  |  |  |  |  |  |
| Total SARS-CoV-2 tests**** | 16,427 | 12,466 | 575 | 2214 | 833 | 83 | 256 |  |
| SARS-CoV-2 | 3,063<br>(18.6) | 2724<br>(21.9) | 40<br>(7.0) | 208<br>(9.4) | 73<br>(8.8) | 3<br>(3.6) | 15<br>(5.9) |  |
| **Seasonal**<br>**distribution,**<br>**n (%)** |  |  |  |  |  |  |  |  |
| Rainy season | 6,528<br>(69.2) | 5,341<sup>a</sup><br>(74.6) | 365<sup>b</sup><br>(46.8) | 387<sup>b</sup><br>(54.2) | 318<sup>b</sup><br>(53.4) | 42<sup>b</sup><br>(76.4) | 75<sup>b</sup><br>(58.1) | <0.001 |

\*The same letters in the same line indicate absence of statistically significant difference.
\*\*The extended PCR panel includes all 8 target viruses, among which are influenza and RSV families. In this table, the extended PCR panel refers to results from adenovirus, endemic coronavirus, hMPV, rhinovirus, and parainfluenza. The total positive tests include detections from both the extended and limited PCR panels and captures the total number of positive cases for influenza and RSV.
\*\*\*Endemic coronavirus (229E, HKU1, NL63, OC43).
\*\*\*\*The total number of tests that includes the SARS-CoV-2 target, collected using both extended and limited PCR panels from 2020 to 2024.

### Viral Detection Distribution

The number of tests conducted was relatively low in the pre-pandemic period (2,099 tests, 5.9% in 2018 and 2019), increased substantially during the pandemic period (25,851 tests, 72.5% in 2020 and 2021) and remained elevated in the post-pandemic period (7,697 tests, 21.6%; Supplementary Table 1). Of the 35,647 tests, 28,955 (81.2%) were performed using an extended PCR panel (Supplementary Table 1). Among these, 8,370 tests were positive (28.9 %), with rhinovirus being the most frequently detected virus (5,024 positive tests, 60.0 %; Table 1). Across all tests, including extended and limited panels, 9,431 (26.5 %) tested positive. The second most common viruses were influenza virus (1,720 positive tests, 18.2%) and RSV (1,395 positive tests, 14.8%).

Other viruses, including hMPV, parainfluenza, adenovirus, and endemic coronaviruses, were detected at lower rates (10.1%, 6.9%, 5.2% and 0.8%, respectively). Of the total 35,647 tests performed, 16,427 included the SARS-CoV-2 target, which was measured from 2020 to 2024 across both the extended and limited panels. SARS-CoV-2 was detected in 3,063 tests (18.6%). Notably, in Curaçao, influenza virus was the predominant pathogen (417 positive tests, 58.4%), differing from the overall trend. Of the 9,431 positive tests, 8,852 (93.9%) were single detections, 527 (5.6%) were co-detections, and 52 (0.5%) involved three or more viruses (Table 1). Given the variation in laboratory testing practices across islands, laboratories, and time periods, testing practices were carefully considered (Supplementary Table 1).

### Seasonality of RTIs in the Caribbean

Seasonal trends of respiratory viral infections are illustrated in Figure 1A. Prior to the COVID-19 pandemic (2018–2020), both rhinovirus and influenza displayed similar seasonal peaks from November to March, with a minor secondary rise around June. During the pandemic years, rhinovirus circulation temporarily decreased but remained detectable year-round, whereas influenza and RSV detections dropped to undetectable levels.

After the pandemic, the prevalence of rhinovirus remained consistently high throughout the year, with recurring peaks in the later months of the year. Influenza, on the other hand, showed an altered pattern in 2022, with a shifted peak between March and December. In subsequent years, however, influenza seasonality appeared to return to pre-pandemic trends, with activity peaking mainly between November and March.

RSV, in contrast to both rhinovirus and influenza, demonstrated a distinct and stable tropical seasonal pattern, with highest prevalence from June to December. Like influenza, RSV activity was undetectable during the pandemic, but its post-pandemic resurgence returned to the previously observed pattern.

GAMs confirmed the statistical significance of these seasonal trends. Rhinovirus showed significant biannual peaks, primarily in November and March (edf = 8.70, χ² = 141.1, p < 0.001; Figure 1B). Influenza demonstrated a clear unimodal seasonal distribution with major peaks from November to March, especially in December and January (edf = 7.96, χ² = 1021.6, p < 0.001; Figure 1C). A minor increase was observed in June, but it did not significantly affect the overall seasonality. RSV activity rose significantly between June and December, with the highest peaks in August and November (edf = 6.64, χ² = 232.1, p < 0.001; Figure 1D).

No significant differences were found between seasonal trends in models that included or excluded the pandemic years, nor between models with and without data from Aruba (Supplementary Figure 1). Patterns were also consistent across age groups (Supplementary Figure 2). Among children, RSV infections increased markedly from June to December (χ² = 181.5, p < 0.001), while among adults the rise occurred slightly later, between August and December (χ² = 73.3, p < 0.001).

Together, these findings highlight that rhinovirus circulation persisted even during pandemic disruptions, while influenza and RSV seasonality proved more vulnerable to external shifts but largely returned to their previous temporal patterns after the pandemic period.

**Figure 1.**
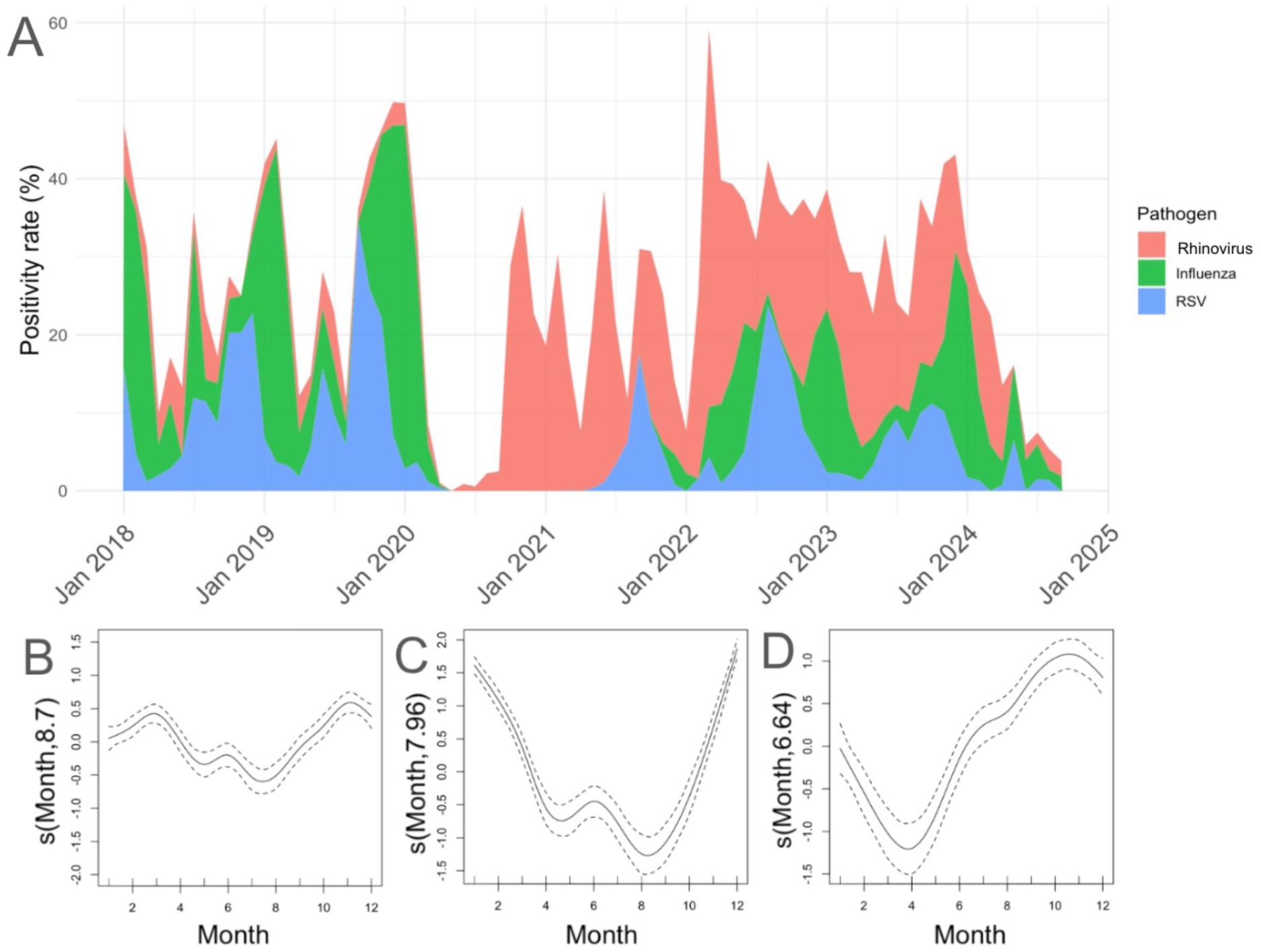
Seasonal Patterns of rhinovirus, influenza virus and RSV in the Caribbean region of the Kingdom of the Netherlands. (A) Positivity rate of rhinovirus (red), influenza virus (green), and RSV (blue) distributed across months and years. (B–D) Partial effect plots for (B) rhinovirus, (C) influenza virus, and (D) RSV by month. The smoothing function *s*(Month) represents the seasonal trend by month, with a 95% confidence interval.

### Impact of Seasonality, Tourism, and host characteristics per Island

Both rhinovirus and influenza demonstrated significant seasonal fluctuations associated with the rainy season, although the direction and magnitude of these effects varied by island (Table 2). Rhinovirus prevalence was significantly higher during the rainy season in Aruba (OR = 1.46; 95% CI: 1.23–1.72; p < 0.001) and Bonaire (OR = 2.49; 95% CI: 1.52-4.19; p < 0.001), and while not significant, the same association was found on Curaçao and St Maarten. Influenza virus was also more frequently detected during the rainy season in Aruba (OR = 2.38 [1.89–2.97]; p < 0.001), but less frequently in Bonaire (OR = 0.07; 95% CI: 0–0.40; p = 0.02) and Sint Maarten (OR = 0.06; 95% CI: 0–0.29; p < 0.01). RSV showed a consistently significant increase during the rainy season, particularly in Aruba (OR = 6.42; 95% CI: 4.26– 9.75; p < 0.001) and Sint Maarten (OR = 7.27; 95% CI: 2.31–28.22; p < 0.01). No significant associations were found for RSV on other islands.

Influenza virus prevalence increased significantly during the high tourism season in Aruba (OR = 8.72; 95% CI: 6.37–12.10; p < 0.001), Curaçao (OR = 3.05; 95% CI: 1.59–6.16; p < 0.01), and Sint Maarten (OR = 10.83; 95% CI: 2.13–198.83; p = 0.02; Table 2). Rhinovirus prevalence was significantly lower during the high tourism season in Aruba (OR = 0.63; 95% CI: 0.54–0.74; p < 0.001), but higher in Sint Maarten (OR = 3.39; 95% CI: 1.54–7.77; p < 0.01). No significant tourism-related associations were observed for influenza or rhinovirus on the other islands, and RSV showed no significant association on any island.

Rhinovirus and RSV showed significantly lower prevalence among adults compared to children, a pattern consistent across Aruba, Curaçao, Sint Maarten, and Bonaire (Table 2) . In contrast, influenza was significantly more prevalent among adults in Curaçao (OR = 2.42; 95% CI: 1.31–4.92; p < 0.01), but significantly less prevalent in adults in Aruba (OR = 0.65; 95% CI: 0.53–0.81; p < 0.001) and Bonaire (OR = 0.38; 95% CI: 0.18–0.87; p = 0.02). No significant age-related associations were observed on other islands, and no significant differences were found by sex.

**Table 2.** Impact of season, tourism and age on respiratory viruses across the Caribbean islands.

|  | Aruba |  | Bonaire |  | Curaçao |  | Sint Maarten* |  |
| --- | --- | --- | --- | --- | --- | --- | --- | --- |
|  | OR<br>[95%CI] | p-value | OR<br>[95%CI] | p-value | OR<br>[95%CI] | p-value | OR<br>[95%CI] | p-value |
| **Rhinovirus** |  |  |  |  |  |  |  |  |
| Climate (rainy season) | 1.46<br>[1.23, 1.72] | <0.001 | 2.49<br>[1.52, 4.19] | <0.001 | 2.14<br>[0.96, 4.88] | 0.07 | 2.49<br>[1.18, 5.28] | 0.02 |
| Tourism (high season) | 0.63<br>[0.54, 0.74] | <0.001 | 1.15<br>[0.68, 1.97] | 0.61 | 0.95<br>[0.44, 2.00] | 0.88 | 3.39<br>[1.54, 7.77] | <0.01 |
| Age group (adults) | 0.37<br>[0.33, 0.43] | <0.001 | 1.35<br>[0.83, 2.19] | 0.22 | 0.39<br>[0.20, 0.72] | <0.01 | 0.17<br>[0.09, 0.31] | <0.001 |
| **Influenza** |  |  |  |  |  |  |  |  |
| Climate (rainy season) | 2.38<br>[1.89, 2.97] | <0.001 | 0.07<br>[0, 0.40] | 0.02 | 1.36<br>[0.79, 2.29] | 0.25 | 0.06<br>[0, 0.29] | <0.01 |
| Tourism (high season) | 8.72<br>[6.37, 12.10] | <0.001 | 3.31<br>[0.53, 67.89] | 0.30 | 3.05<br>[1.59, 6.16] | <0.01 | 10.83<br>[2.13, 198.83] | 0.02 |
| Age group (adults) | 0.65<br>[0.53, 0.81] | <0.001 | 0.38<br>[0.18, 0.87] | 0.02 | 2.42<br>[1.31, 4.92] | <0.01 | 0.55<br>[0.29, 1.04] | 0.06 |
| **RSV** |  |  |  |  |  |  |  |  |
| Climate (rainy season) | 6.42<br>[4.26, 9.75] | <0.001 | 0.62<br>[0.05, 4.77] | 0.68 | 1.85<br>[0.67, 5.06] | 0.23 | 7.27<br>[2.31, 28.22] | <0.01 |
| Tourism (high season) | 1.45<br>[1, 2.09] | 0.05 | 0.88<br>[0.11, 9.88] | 0.91 | 0.92<br>[0.34, 2.45] | 0.87 | 0.60<br>[0.21, 1.58] | 0.31 |
| Age group (adults) | 0.04<br>[0.03, 0.06] | <0.001 | 0.12<br>[0.02, 0.47] | <0.01 | 0.30<br>[0.13, 0.68] | <0.01 | 0.02<br>[0, 0.10] | <0.001 |

\*Sint Maarten cohort combined with Saba and Sint Eustatius

### Clinical Burden of RTIs in the Caribbean

A total of 1,521 tests were conducted across Sint Maarten, Saba, and Sint Eustatius, including 788 episodes involving patients admitted to the SMMC with RTI symptoms. After applying predefined exclusion criteria, 593 patients were included in the clinical cohort. Of these, 235 patients (39.6%) tested positive for a viral infection (Table 3).

Rhinovirus was the most frequently detected pathogen (n = 132, 56.2%), followed by influenza virus (n = 36, 15.3%) and RSV (n = 33, 14.0%). Other viruses such as, adenovirus, endemic coronavirus, hMPV and parainfluenza virus tested positive in the remaining 34 patients. Co-infections were observed in 51 cases (21.7%), most commonly rhinovirus in combination with RSV (n = 12, 5.1%) or adenovirus (n = 14, 6.0%). The cohort was predominantly paediatric, (n= 421, 71.0%) with a median age of 4 years (IQR: 2– 42).

Female patients made up a slight majority (n = 326, 55.0%). Rhinovirus and RSV were predominantly detected in children (n = 114, 86.4% and n = 28, 84.8%, respectively), whereas influenza virus was more frequently identified in adults (n = 20, 55.6%). The most frequently reported symptoms among virus-positive cases were coughing/sneezing (n = 155, 66.0%) and fever (n = 119, 50.6%) (Table 3).

**Table 3.**
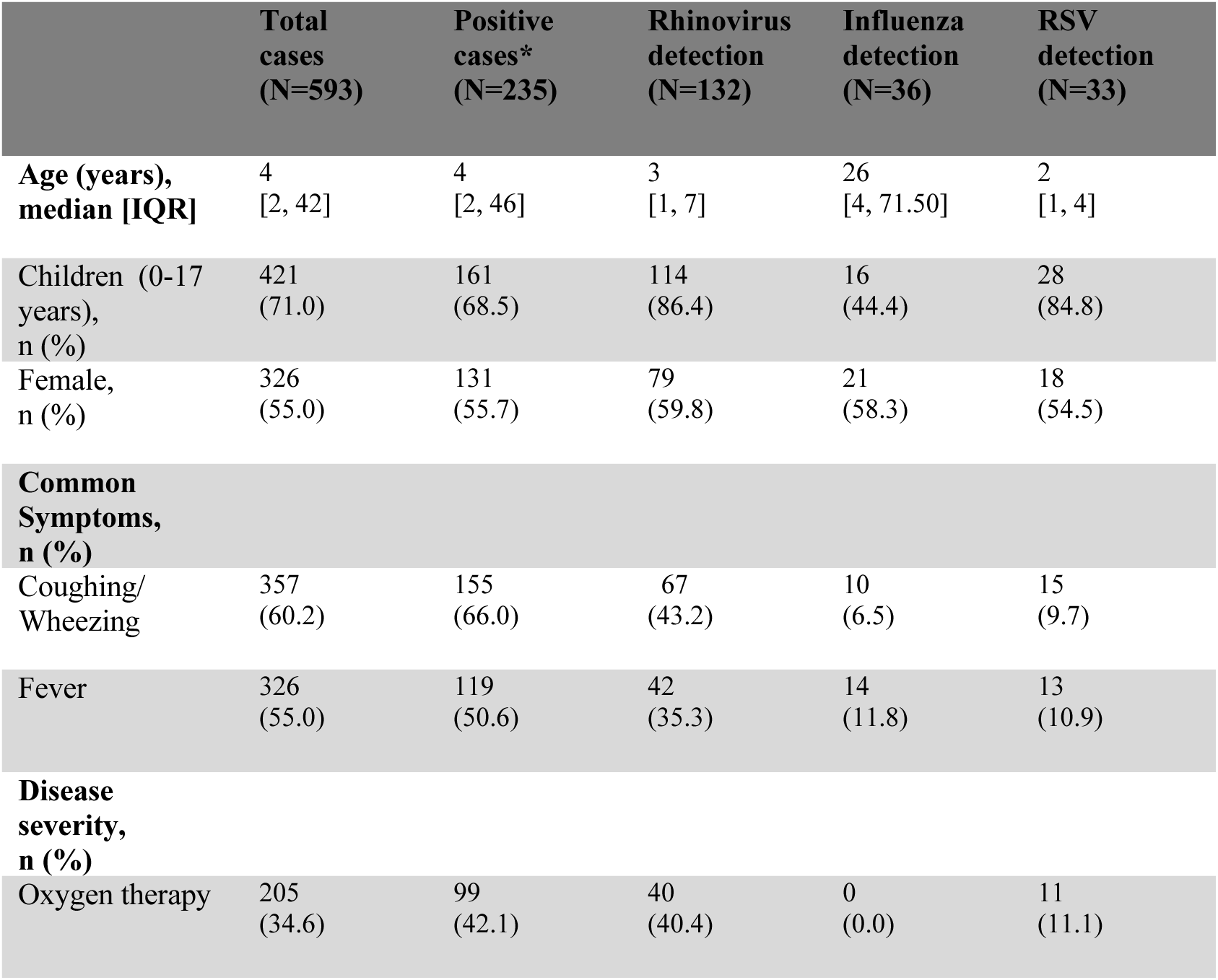
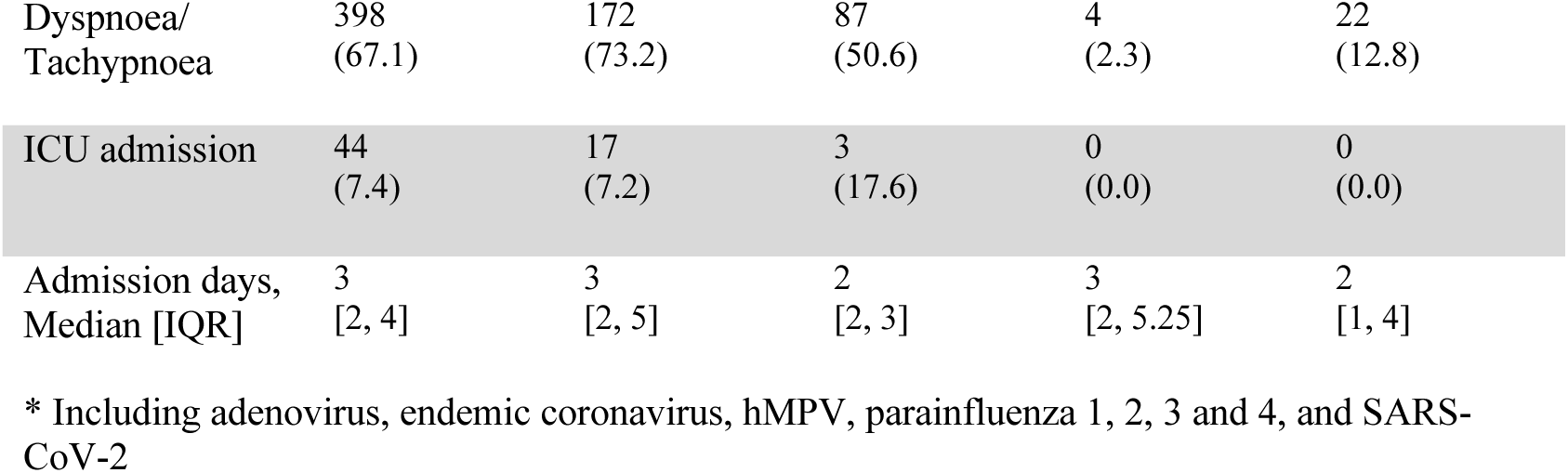
Baseline characteristics of the clinical cohort.

|  | **Total cases**<br>**(N=593)** | **Positive cases\***<br>**(N=235)** | **Rhinovirus detection**<br>**(N=132)** | **Influenza detection**<br>**(N=36)** | **RSV detection**<br>**(N=33)** |
| --- | --- | --- | --- | --- | --- |
| **Age (years), median [IQR]** | 4<br>[2, 42] | 4<br>[2, 46] | 3<br>[1, 7] | 26<br>[4, 71.50] | 2<br>[1, 4] |
| **Children (0-17 years), n (%)** | 421<br>(71.0) | 161<br>(68.5) | 114<br>(86.4) | 16<br>(44.4) | 28<br>(84.8) |
| **Female, n (%)** | 326<br>(55.0) | 131<br>(55.7) | 79<br>(59.8) | 21<br>(58.3) | 18<br>(54.5) |
| **Common Symptoms, n (%)** |  |  |  |  |  |
| **Coughing/ Wheezing** | 357<br>(60.2) | 155<br>(66.0) | 67<br>(43.2) | 10<br>(6.5) | 15<br>(9.7) |
| **Fever** | 326<br>(55.0) | 119<br>(50.6) | 42<br>(35.3) | 14<br>(11.8) | 13<br>(10.9) |
| **Disease severity, n (%)** |  |  |  |  |  |
| **Oxygen therapy** | 205<br>(34.6) | 99<br>(42.1) | 40<br>(40.4) | 0<br>(0.0) | 11<br>(11.1) |
| Dyspnoea/<br>Tachypnoea | 398<br>(67.1) | 172<br>(73.2) | 87<br>(50.6) | 4<br>(2.3) | 22<br>(12.8) |
| ICU admission | 44<br>(7.4) | 17<br>(7.2) | 3<br>(17.6) | 0<br>(0.0) | 0<br>(0.0) |
| Admission days,<br>Median [IQR] | 3<br>[2, 4] | 3<br>[2, 5] | 2<br>[2, 3] | 3<br>[2, 5.25] | 2<br>[1, 4] |

\* Including adenovirus, endemic coronavirus, hMPV, parainfluenza 1, 2, 3 and 4, and SARS-CoV-2

Rhinovirus detection was significantly associated with the need for oxygen therapy compared to non-rhinovirus cases (OR = 2.78, [1.72-4.50]; p < 0.001; Table 4). Adults were more likely than children to require oxygen therapy (OR = 7.39, [4.70-11.78]; p < 0.001), and each additional day of hospitalization was associated with an increased likelihood of oxygen therapy (OR = 1.25, [1.17-1.37]; p < 0.001). Dyspnoea or tachypnoea was also significantly more prevalent in patients with rhinovirus infection (OR = 2.26, [1.42-3.67]; p < 0.001; Table 4), and in adults compared to children (OR = 3.18, [1.99-5.22]; p < 0.001). Symptoms occurred more frequently during the rainy season (OR = 1.65, [1.15-2.38]; p < 0.01). There were no statistically significant differences in the length of hospital stay between patients infected with different viruses (Supplementary Figure 3). Among the 205 patients who required oxygen therapy, 22 (10.7%) had co-infections involving rhinovirus and either RSV or adenovirus. Similarly, of the 398 patients experiencing dyspnoea or tachypnoea, 38 (9.5%) had co-infections with rhinovirus and RSV or adenovirus.

**Table 4.**
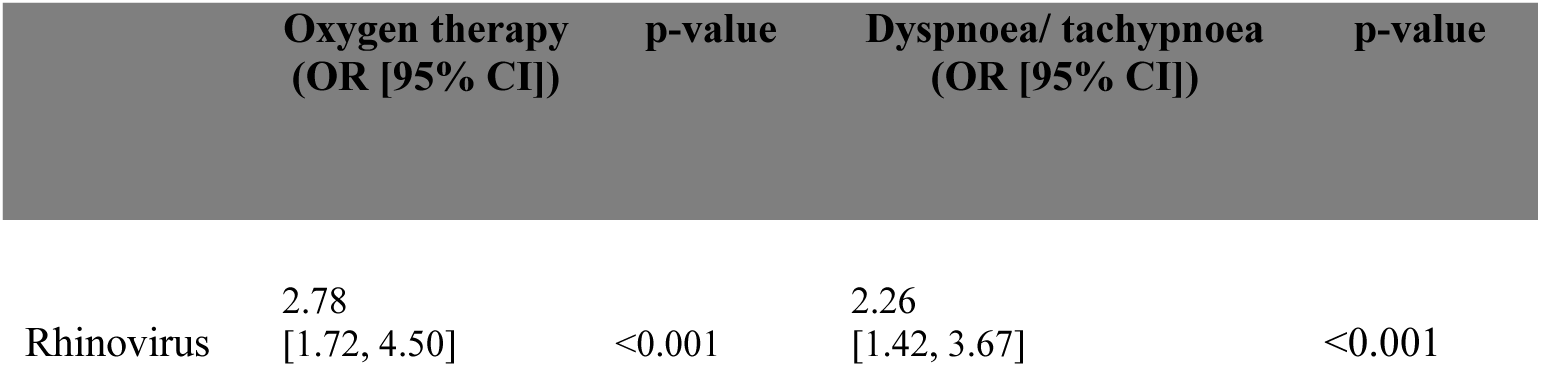
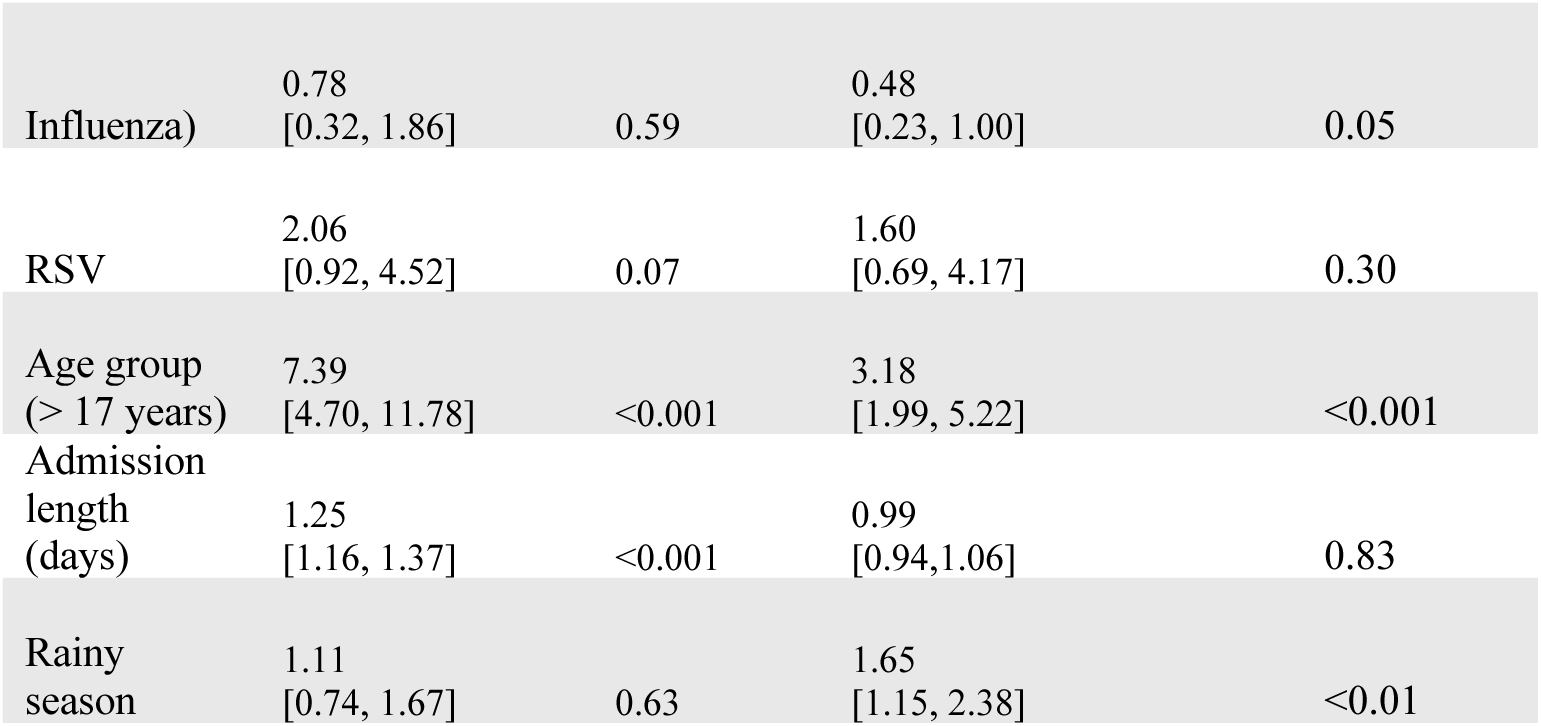
Disease severity of RTIs in hospitalised patients.

|  | Oxygen therapy<br>(OR [95% CI]) | p-value | Dyspnoea/ tachypnoea<br>(OR [95% CI]) | p-value |
| --- | --- | --- | --- | --- |
| Rhinovirus | 2.78<br>[1.72, 4.50] | <0.001 | 2.26<br>[1.42, 3.67] | <0.001 |
| Influenza) | 0.78<br>[0.32, 1.86] | 0.59 | 0.48<br>[0.23, 1.00] | 0.05 |
| RSV | 2.06<br>[0.92, 4.52] | 0.07 | 1.60<br>[0.69, 4.17] | 0.30 |
| Age group<br>(> 17 years) | 7.39<br>[4.70, 11.78] | <0.001 | 3.18<br>[1.99, 5.22] | <0.001 |
| Admission<br>length<br>(days) | 1.25<br>[1.16, 1.37] | <0.001 | 0.99<br>[0.94, 1.06] | 0.83 |
| Rainy<br>season | 1.11<br>[0.74, 1.67] | 0.63 | 1.65<br>[1.15, 2.38] | <0.01 |

## Discussion

### Key Findings and Public Health Implications

This is the first study to comprehensively describe the seasonal patterns and clinical burden of respiratory viral infections across all six Caribbean islands of the Kingdom of the Netherlands, filling a critical knowledge gap in tropical respiratory epidemiology. In alignment with WHO recommendations to tailor preventive strategies to local epidemiology^11,12^, our findings offer important guidance for optimizing the timing of interventions such as influenza vaccination and RSV prophylaxis. Notably, influenza activity peaked between November and March, aligning with temperate-zone seasonality rather than the year-round circulation often reported in tropical settings.^28,29^ In contrast, RSV followed a more expected tropical pattern^29,30^, with peaks during the rainy season, yet current prophylaxis policies across the islands remain based on Dutch schedules, resulting in misalignment with local circulation patterns. Furthermore, rhinovirus emerged as the most frequently detected virus across all age groups and was significantly associated with hospitalization, oxygen requirement, and respiratory distress, especially in young children.

These findings highlight a substantial paediatric disease burden in the absence of vaccines or monoclonal therapies and underscore an urgent need for follow-up studies into rhinovirus epidemiology, environmental drivers, and potential interactions with co-circulating pathogens.

### Seasonal Drivers: Climate, Tourism, and Behaviour

Our data show that influenza activity peaked between November and March. Notably, coinciding with the high tourist seasons on Aruba, Curaçao, and Sint Maarten, suggesting that human mobility, crowding, and widespread use of air-conditioned indoor environments may play a more prominent role than rainfall in driving transmission. Consequently, islands like Aruba, Curaçao, and Sint Maarten, with relatively small resident populations but substantial year-round tourism, may face amplified transmission due to increased crowding and population movement.^17,20,28,31–33^ In contrast, smaller islands like Saba and Sint Eustatius, which receive fewer tourists and have more dispersed populations, may experience lower crowding, potentially limiting the spread of these viruses.

RSV showed strong seasonality aligned with the rainy season (June–December), supporting prior research indicating that RSV transmission in tropical regions, such as the Caribbean, Southeast Asia, and Sub-Saharan Africa, is facilitated by high humidity, rainfall, and increased indoor crowding during the rainy season.^13,28–32^ Rhinovirus circulated year-round and was less influenced by tourism or seasonal factors, pointing to high environmental stability due to its non-enveloped structure.^30,34^

### Clinical Burden and Paediatric Impact

In our clinical cohort from the Windward Islands, rhinovirus was the dominant pathogen among hospitalised patients and showed strong associations with the need for oxygen therapy and signs of respiratory distress. Notably, this sub-analysis confirmed rhinovirus as the only respiratory virus significantly linked to both clinical severity and supportive care needs. Co-detections with RSV and adenovirus were frequent and may exacerbate disease severity, especially in vulnerable paediatric populations.^13,35,36^ These observations reinforce the need for paediatric capacity planning that accounts for high rhinovirus circulation, even in the absence of specific prevention tools.

### Disparities Across Islands: Health Systems and Access

Although all six islands are part of the Kingdom of the Netherlands, substantial differences exist in healthcare access and infrastructure. The BES islands (Bonaire, Saba, and Sint Eustatius), which are directly governed by the Netherlands, benefit from automatic inclusion in national health programs, including immunization schedules. In contrast, the CAS islands (Curaçao, Aruba, and Sint Maarten) maintain autonomous healthcare systems and must independently coordinate implementation and funding of new public health interventions, often relying on partnerships with organisations such as the Pan American Health Organization (PAHO). Laboratory capacity also varied widely across the region: Aruba contributed the majority of virological data, while smaller islands depended on referral centres such as SMMC in Sint Maarten.

### Strengths, Limitations, and Future Directions

This study leverages a unique multi-island dataset collected over multiple years, enabling robust statistical modelling of virus seasonality and clinical severity. However, limitations include heterogeneity in testing intensity across islands, incomplete data during the COVID-19 pandemic, and underrepresentation of mild or non-hospitalised cases. Still, these findings provide a critical empirical basis to inform regional public health policy.

Climate-related shifts in temperature, rainfall, and extreme weather events are expected to increasingly influence respiratory virus circulation in tropical settings. These changes may alter seasonality, intensify transmission, and raise exposure risks, particularly for young children.^37–40^ Our data show that RSV seasonality differs significantly from that of the European Netherlands, while prophylaxis policies on the islands are still modelled after Dutch guidelines. Aligning the timing of RSV immunization, whether through nirsevimab or maternal pre-fusion vaccination^41,42^, with local circulation patterns will be crucial to reducing early infant morbidity. For the BES islands, implementation is expected to be centrally coordinated by the Dutch Ministry of Health, while the CAS islands will require independent planning. As a result, rollout remains uncertain. In contrast, influenza vaccination timing appears largely appropriate, as Caribbean peaks roughly align with those in the Netherlands; however, this alignment was previously assumed rather than empirically validated, underscoring the value of locally derived data.

Lastly, the potential for co-infection between respiratory and arboviral pathogens warrants further exploration. Arboviruses such as dengue and Zika circulate year-round in the region^22^, and recent studies suggest that viral co-infections, such as SARS-CoV-2 and dengue, can increase morbidity and mortality.^43^ Although our dataset did not include arboviral diagnostics, it is plausible that co-infection or co-endemicity may have contributed to the clinical severity observed with rhinovirus. This hypothesis, along with the virus’s high environmental persistence, should be investigated further in prospective studies. Such insights are essential to improve syndromic surveillance, inform integrated preparedness strategies, and tailor clinical management to the unique challenges of tropical island health systems.

## Conclusion

These findings underscore the urgency of implementing interventions that are tailored to local viral patterns and healthcare capacities, informed by climate realities, and designed to reduce inequities between islands, ensuring that all populations, regardless of geography or governance structure, have access to timely and effective respiratory disease prevention, while also highlighting the need for strengthened surveillance, adaptive immunization strategies, and continued research into evolving respiratory threats.

## Acknowledgements

We thank all participants, nurses, laboratory technicians, hospitals and laboratories that contributed to this study and provided data.

## Ethics statement

This study used fully anonymised laboratory surveillance data, with no personal identifier or individual-level analysis.

The Ethics Committee of SMMC approved this study and the clinical data collection, providing a waiver of consent.

## Patient and Public Involvement statement

Patients or the public were not involved in the design, conduct, reporting, or dissemination plans of this research.

## Data Availability Statement

The datasets generated and analysed in this study are not publicly available due to institutional data sharing agreements. However, these data are available from the corresponding author upon reasonable request, solely for the purposes of peer review.

## Author Reflexivity Statement

This study was conducted through collaboration between researchers based in both European and Caribbean institutions. We acknowledge potential imbalances in research infrastructure and resources and strived for equitable partnership throughout the study. Authors from both regions collaboratively contributed to the design, analysis, and manuscript development.

## List of abbreviations

ANOVA: Analysis of Variance
BES islands: Bonaire, Saba, and Sint Eustatius
CAS islands: Curaçao, Aruba, and Sint Maarten
COVID-19: Coronavirus disease 2019
Edf: Effective degrees of freedom
GAMs: Generalised Additive Models
GLMs: Generalised Linear Models
hMPV: human metapneumovirus
ICU: Intensive Care Unit
LRTIs: Lower Respiratory Tract Infections
PAHO: Pan American Health Organization
PCR: Polymerase Chain Reaction
RSV: Respiratory syncytial virus
RTIs: Respiratory Tract Infections
SARS-CoV-2: Severe Acute Respiratory Syndrome Coronavirus 2
SMMC: Sint Maarten Medical Center
WHO: World Health Organization

