## Supplementary Materials for "Seasonal Dynamics of Influenza and RSV in the Caribbean: A Call for Regionally Tailored Preventive Measures"

### Supplementary tables and figures

#### Table 1. Laboratory testing practices on the six Caribbean islands

| Laboratory testing practices | Aruba | Bonaire | Curaçao | Sint Maarten | Saba | Sint Eustatius | Total |
|---|---|---|---|---|---|---|---|
| Pre-pandemic (2018-2019) | 1,455 (4.9) | 165 (13.1) | 358 (10.9) | 74 (6.9) | 23 (15.6) | 24 (7.9) | 2,099 (5.9) |
| During-pandemic (2020-2021) | 24,083 (81.4) | 562 (44.8) | 827 (25.2) | 272 (25.5) | 73 (49.7) | 34 (11.1) | 25,851 (72.5) |
| Post-pandemic (2022-2024) | 4,051 (13.7) | 528 (42.1) | 2,099 (63.9) | 721 (67.6) | 51 (34.7) | 247 (81.0) | 7,697 (21.6) |
| **Testing panels** | | | | | | | |
| Extended panels*, n (%) | 25,237 (85.3) | 1,255 (100) | 1,055 (32.1) | 1,067 (100) | 147 (100) | 194 (63.6) | 28,955 (81.2) |
| Limited panels**, n (%) | 4,352 (14.7) | 0 (0) | 2,229 (67.9) | 0 (0) | 0 (0) | 111 (36.4) | 6,692 (18.8) |

*Extended panels including all seven viruses
**Limited panels including influenza and RSV

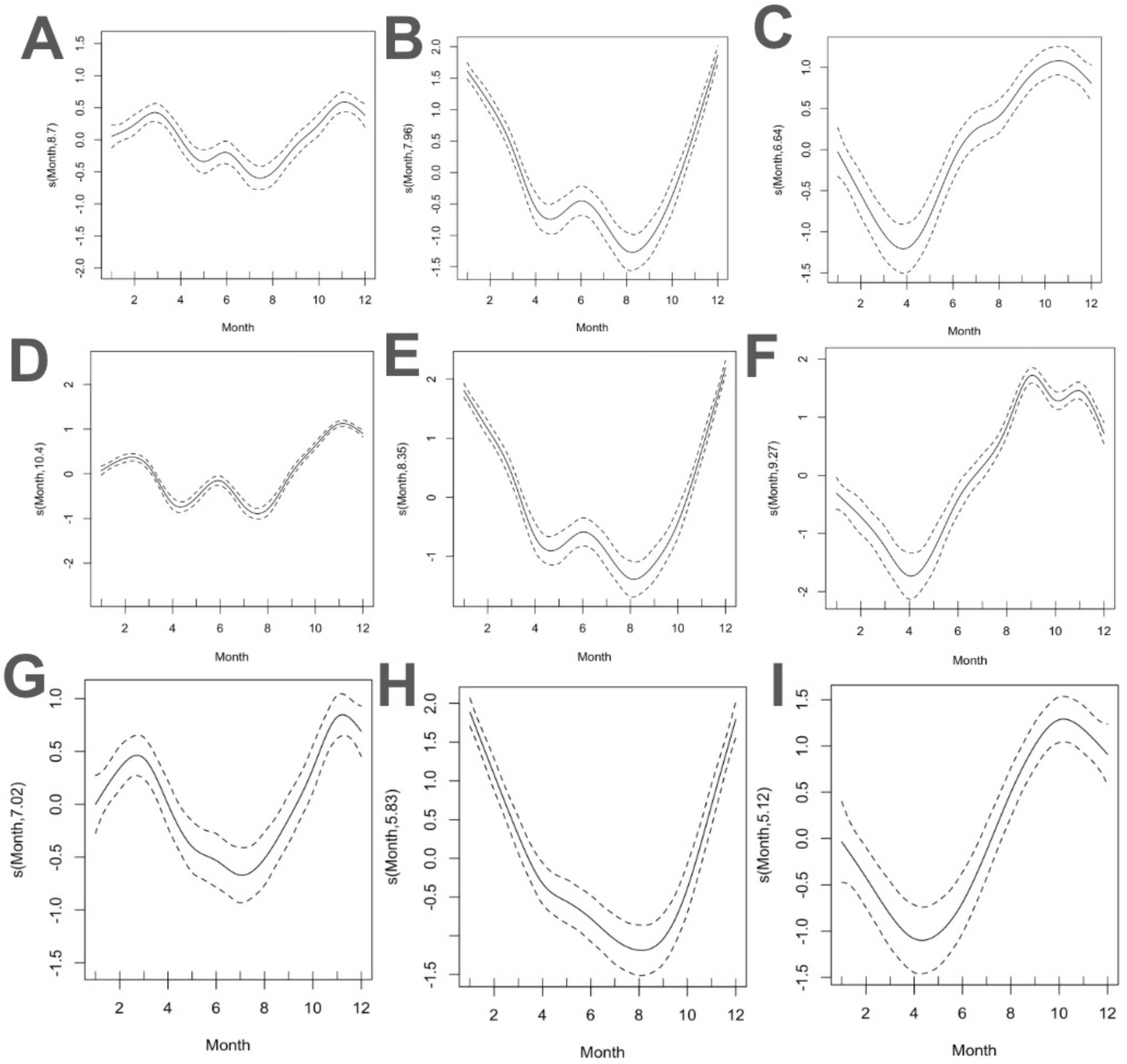


**Figure 1. Seasonal Patterns of Rhinovirus, Influenza and RSV in the Caribbean region of the Kingdom of the Netherlands.** (A–D) Partial effect plots for (A) rhinovirus, (B) influenza, and (C) RSV seasonality without the pandemic period (2020-2021). (D–F) Partial effect plots for (D) rhinovirus, (E) influenza, and (F) RSV seasonality from 2018 to 2024. (G–I) Partial effect plots for (G) rhinovirus, (H) influenza, and (I) RSV seasonality without data from Aruba and pandemic period (2020-2021). The smoothing function *s*(Month) represents the seasonal trend by month, with a 95% confidence interval.

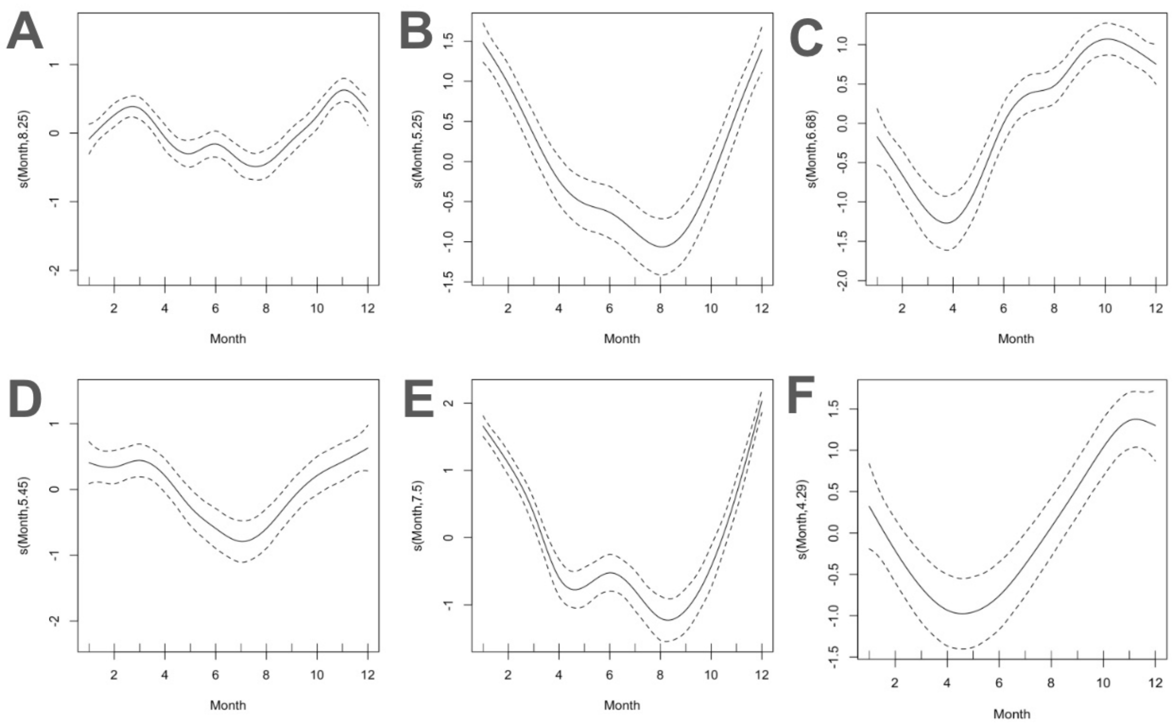


**Figure 2. Seasonal trend of viral infections for children and adults.** (A-C) Partial effect plots for (A) rhinovirus, (B) influenza, and (C) RSV by month for the children population, and (D-F) partial effect plots for (D) rhinovirus, (E) influenza, and (F) RSV by month for the adult population.

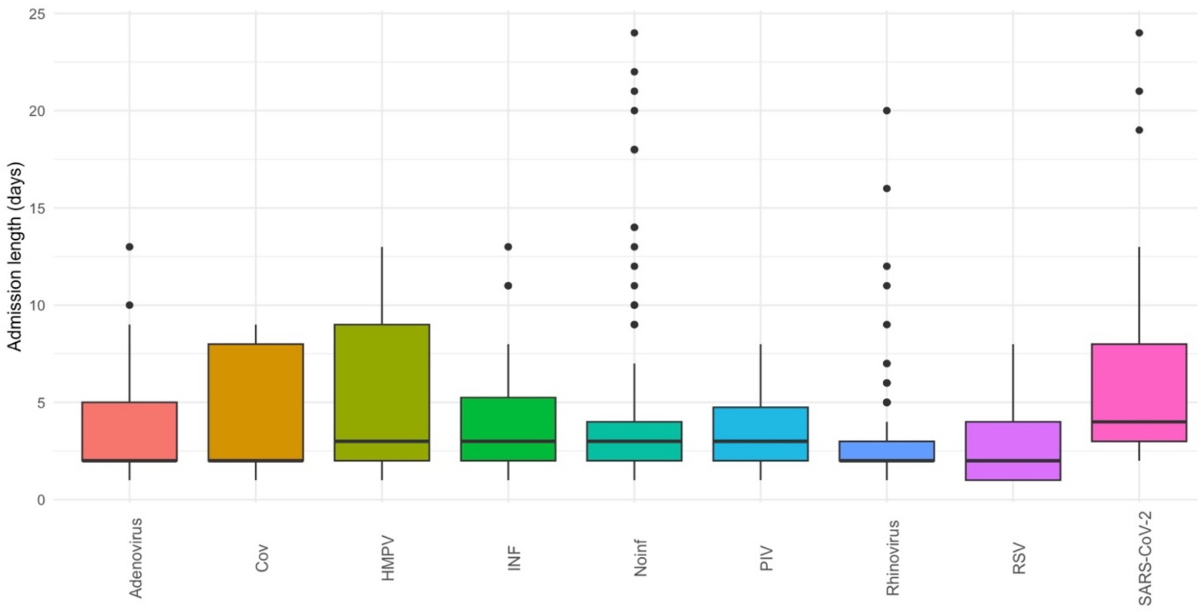


**Figure 3. Distribution of length of admission in hospital according to detected viral infections.**
Admission length of adenovirus, endemic coronavirus (CoV), human metapneumovirus (HMPV), influenza virus (INF), no infections (Noinf), parainfluenza (PIV), rhinovirus, RSV, and SARS-CoV2 were displayed.
